# Evaluation of a Commercially Available Rapid RT-PCR Assay’s Detection of SARS-CoV-2 Novel Variants

**DOI:** 10.1101/2023.01.30.23285171

**Authors:** Lewis Back, Jennifer K. Logue, Caitlin R. Wolf, Peter D. Han, Lea M. Starita, Helen Y. Chu

**Affiliations:** Division of Allergy and Infectious Diseases, Department of Medicine, University of Washington, Seattle (98195), Washington, USA; Brotman Baty Institute for Precision Medicine, Seattle (98195), Washington, USA; Department of Genome Sciences, University of Washington, Seattle (98195), Washington, USA

## Abstract

Rapid molecular diagnostic tests have been critical in the response to the COVID-19 pandemic. It is important to evaluate the ability of these assays to identify variants of concern, particularly across varying collection and storage conditions. Nasal swabs positive for Alpha (B.1.1.7), Beta (B.1.351), Delta (B.1.167.2), Gamma (P.1), or Omicron (B.1.1.529) variants of concern (VOCs) were stored in TE buffer and viral transport media (VTM). We evaluated the sensitivity of the Cepheid Xpert® Xpress SARS-CoV-2 assay in detecting VOC samples and validated TE buffer for use with the assay. Testing of known VOC positives revealed no substantial reduction of PCR sensitivity. Comparison of TE and VTM samples also revealed no reduction in performance when using TE buffer, validating the use of TE buffer to store SARS-CoV-2 samples. SARS-CoV-2 VOCs collected and stored across various conditions can be detected by the Cepheid Xpert® Xpress SARS-CoV-2 assay.

## Background

Throughout the COVID-19 pandemic, rapid and accurate diagnosis of Severe Acute Respiratory Syndrome Coronavirus 2 (SARS-CoV-2) infection has been critical to managing viral spread. Reverse transcriptase-polymerase chain reaction (RT-PCR) tests for detecting SARS-CoV-2 were some of the first diagnostic tools to be developed during the global pandemic and became a pivotal technology in diagnosing SARS-CoV-2 infection worldwide. The Xpert® Xpress SARS-CoV-2 assay (Cepheid) for point-of-care testing was among the earliest RT-PCR assays available, having obtained EUA status in March 2020 [1]. The assay is marketed as a rapid on-demand test that can accurately detect the presence of SARS-CoV-2 in under an hour using primers that target the viral envelope (*E*) and nucleocapsid (*N2*) genes.

The Xpert® Xpress SARS-CoV-2 assay has shown concordance with the Cobas® SARS-CoV-2 assay (Roche) and the Centers for Disease Control and Prevention (CDC) RT-PCR test, and increased sensitivity compared to the Abbott ID Now SARS-CoV-2 Assay [2,3,4]. However, since the assay’s development using the USA-WA1/2020 strain, numerous novel genetic variants have been observed circulating in the population. As of January 2023, the Omicron variant, which contains more than 30 mutations in the spike protein-coding region of its genome, is estimated to account for nearly all coronavirus variants currently circulating in the United States [5]. Variants can affect performance of diagnostic PCR assays due to mutations in primer binding sites that may lead to false negative results. Reduced sensitivity and amplification of the *S*-gene target in variant samples, also known as *S*-gene target failure, has been observed in several EUA-authorized tests for SARS-CoV-2 [6]. Assays that target other genes, such as the Xpert® Xpress, may be able to detect those variants with *S*-gene mutations that might otherwise cause target failure. However, several single nucleotide polymorphisms (SNPs) within the nucleocapsid gene have also been reported to cause *N2*-gene target failure in the Xpert® Xpress assay [7].

### Objectives

Given the increased burden of COVID-19 due to VOCs and the potential impact on testing performance, we assessed whether the Xpert® Xpress SARS-CoV-2 assay could detect VOCs without substantial reductions in sensitivity, by comparing results from the Xpert® Xpress assay to those from a clinically certified RT-PCR assay.

The tested VOC samples were stored in low TE (Tris EDTA) buffer, a medium which has not been formally evaluated for use with the Xpert® Xpress. Prior to testing the VOC samples, we validated the use of TE buffer as a storage medium. SARS-CoV-2 positive controls stored in TE buffer were run on the Xpert® Xpress and compared to positive controls stored in standard viral transport medium (VTM; Becton, Dickinson and Company, Franklin, New Jersey, USA).

### Study Design

To verify the use of low TE buffer (10 mM Tris-HCl pH 7.5, 0.1 mM EDTA) as a storage medium, 19 positive controls were prepared using Microbiologics HE0065 lyophilized SARS-CoV-2 pellets. 10 samples were prepared in TE buffer and 9 samples were prepared in VTM. The pellets were hydrated per manufacturer specifications by vortexing in provided 1.5 mL DI H_2_O tubes [8]. 100 μL of rehydrated whole virus was pipetted into each 3mL tube containing either TE buffer or VTM and vortexed again. Samples were stored at -80°C until use. All samples were loaded into Xpert® Xpress cartridges using the provided fixed-volume 300 μL pipettes under aseptic conditions in a biosafety cabinet and were tested per Cepheid protocols [9].

To assess the assay’s detection of VOCs, we tested 46 SARS-CoV-2 positive anterior nasal swabs stored in TE buffer. Twenty-nine Alpha (B.1.1.7), 5 Beta (B.1.351), 4 Delta (B.1.167.2), and 7 Gamma (P.1) samples were tested with the Xpert® Xpress SARS-CoV-2 assay. Six Omicron (B.1.1.529) samples were tested with the Xpert® Xpress SARS-CoV-2/Flu/RSV assay due to inventory availability at the time of testing. The single plex assay contained separate channels for *E* and *N2* genes while the triplex assay contained one combined channel for *E* and *N2* genes.

The VOC samples were collected in Seattle, Washington from March 2021 through December 2021. All samples were previously tested and determined positive for SARS-CoV-2 by the Brotman Baty Institute’s (BBI) Advanced Technology Lab (BATLab), using a laboratory-developed, clinically certified RT-qPCR test consisting of 2 replicate assays each for the *ORF1b*-gene and *S*-gene, and 4 multiplex assays for human marker RNase P [10]. A positive result was indicated if 3-4 reactions for RNase P had cycle threshold (Ct) values earlier than 36 cycles, and 2-4 SARS-CoV->2 wells had C_t_ values earlier than 40 cycles. Samples were subsequently sequenced using the Illumina COVIDSeq Kit followed by genome assembly on a modified iVar pipeline [11]. Variants were then identified using the Pangolin Web App [12].

## Results

Of the 9 VTM positive controls tested, 8 returned positive and 1 returned negative. All 10 TE buffer controls tested returned positive (**Figure 1**). Upon testing samples of all five variants, we found the Cepheid rapid RT-PCR results to be concordant with the original BBI RT-PCR assay, with a 100% sensitivity among Beta, Delta, Gamma, and Omicron samples and a 96.55% positivity rate among Alpha samples. One instrument error occurred while running Alpha samples which caused the discrepancy in positivity. Omitting the instrument error raised the positivity of Alpha samples to 100% (**Figure 2**).

**Figure 1.**
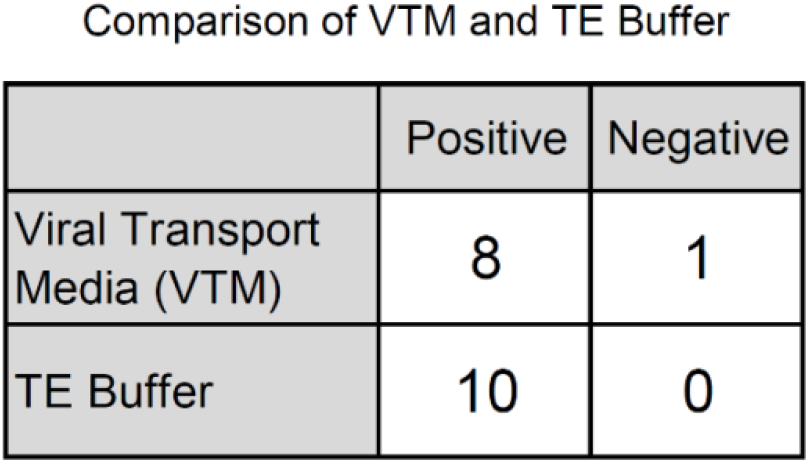
Results of TE Buffer and VTM controls on rapid RT-PCR assay. Positive control samples contained in viral transport medium (VTM) and TE buffer were run on the Xpert® Xpress assay to validate the use of TE buffer as a medium. Positive and negative control values for each medium are listed.

**Figure 2.**
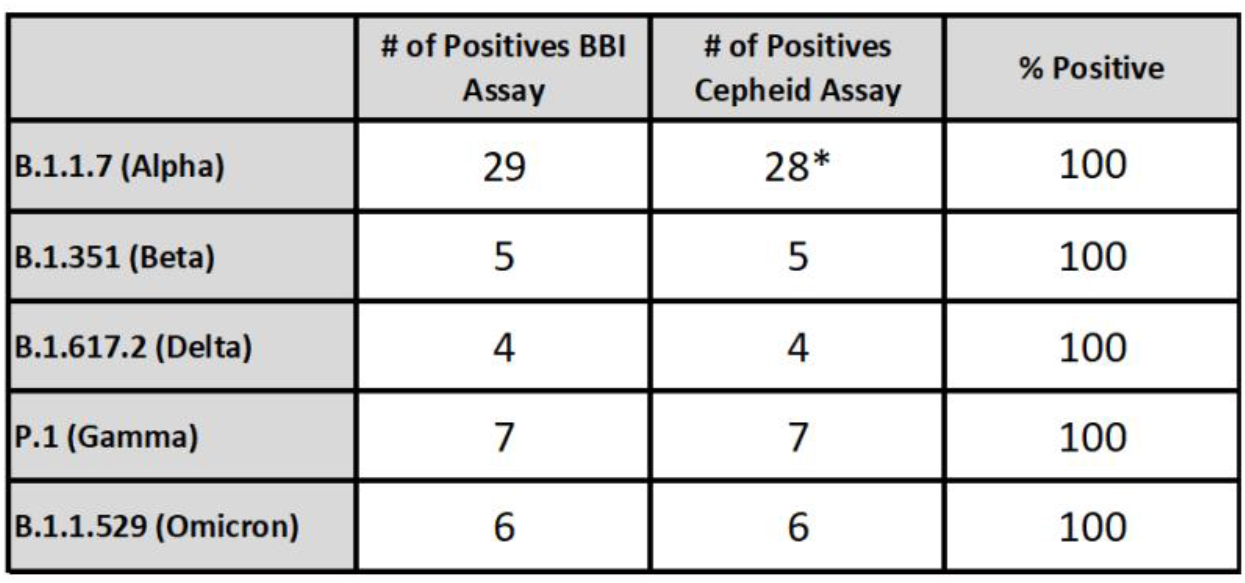
Number of positive results for both original and rapid RT-PCR assays and percent positivity of rapid RT-PCR results, separated by variant of concern.Percent positivity was calculated by dividing Cepheid assay positive results from BBI assay positive results. *One instrument error occurred during testing. Percent positivity for Alpha samples was adjusted to 100% after omitting the instrument error.

The C_t_ values for BBI RT-PCR assay showed substantial reduction in *S*-gene amplification for specimens with the Delta variant (*S*-gene target failure) (**Figure 3a**). This is expected as the primer binding site for the laboratory developed *S*-gene assay is disrupted by the Delta Δ156-157 mutation. The Cepheid assay, however, showed no substantial reduction of amplification in *E* or *N2* genes (**Figure 3b**).

**Figures 3a and 3b.**
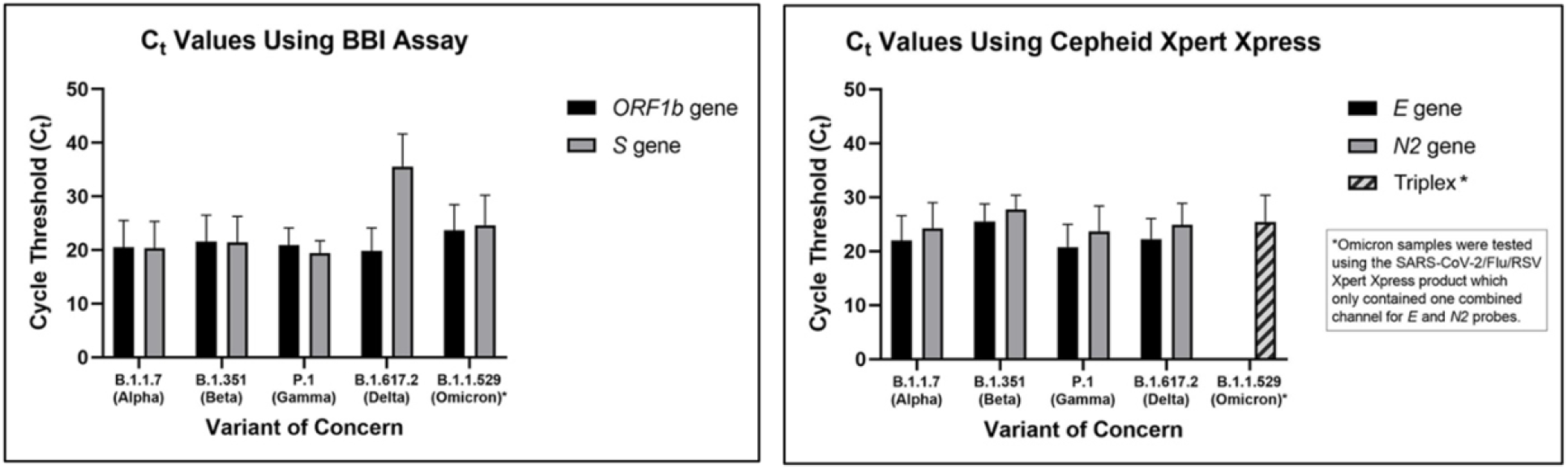
Average C_t_ value results from both original assay (3a, left) and rapid RT-PCR assay (3b, right) grouped by variant of concern. Error bars represent one standard deviation

## Discussion

Many diagnostic tests have been developed during the COVID-19 pandemic; however, manufacturing and supply shortages, due to the immense burden of SARS-CoV-2 infections worldwide, have led to new methods of sample collection and storage. Insufficient supply in the standard collection media, VTM, prompted the use of other storage mediums, such as low TE Buffer, that were not originally validated for use with many of the original diagnostic assays. The results of this study indicate that testing of SARS-CoV-2 samples stored in low TE buffer is concordant to VTM and does not diminish sensitivity of rapid molecular assays.

Newly emerging strains of SARS-CoV-2 have increased the spread of COVID-19 through various mechanisms, including evading detection by RT-PCR. The Omicron variant is a prime example of a VOC with potentially significant *S*-gene target failure due to its unusually high number of *S*-gene mutations. Our results indicate that the Xpert® Xpress SARS-CoV-2 assays are able to accurately detect SARS-CoV-2 variants despite high numbers of mutations in the viral genome. It is likely that other molecular assays that do not target the *S*-gene will demonstrate accurate detection of the SARS-CoV-2 variants tested here, including Omicron and its subvariants with numerous mutations in the *S*-gene region. However, only the B.1.1.529 variant was tested during this study, as it was the only widely circulating Omicron variant at the time of testing. Additionally, no appreciable *N2*-gene target failure was observed in the Xpert® Xpress; for all samples, the *N2* gene was detected with C_t_ values lower than 34.6. Other assays should be validated for diagnostic use in identifying infections with SARS-CoV-2 variants and further testing of the Xpert® Xpress SARS-CoV-2 is required in the case of variants with numerous *E* or *N2* gene mutations.

Rapid RT-PCR is crucial in quickly identifying and isolating infected individuals and this holds especially true for novel variants of concern. This work showed good correlation and diagnostic accuracy of the commercially available Xpert® Xpress system, compared to a clinically certified RT-PCR assay.

## Data Availability

All data produced in the present study are available upon reasonable request to the authors.

## Declaration of Competing Interest

HYC reported consulting with Ellume, Pfizer, The Bill and Melinda Gates Foundation, Glaxo Smith Kline, and Merck. She has received research funding from Emergent Ventures, Gates Ventures, Sanofi Pasteur, The Bill and Melinda Gates Foundation, and support and reagents from Ellume and Cepheid outside of the submitted work.

## Funding

Cepheid provided cartridges free of charge. However, Cepheid did not contribute to sample and data collection, analysis, or interpretation.

## Notes

### Author Declarations

The University of Washington Institutional Review Board gave ethical approval for this work.

## References

1. Hinton, D. (2021, January 7). Xpert Xpress SARS-CoV-2—Letter of Authorization. https://www.fda.gov/media/136316/download

2. Goldenberger, D., Leuzinger, K., Sogaard, K. K., Gosert, R., Roloff, T., Naegele, K., Cuénod, A., Mari, A., Seth-Smith, H., Rentsch, K., Hinić, V., Hirsch, H. H., & Egli, A. (2020). Brief validation of the novel Xpert® Xpress SARS-CoV-2 PCR assay. Journal of Virological Methods, 284, 113925. https://doi.org/10.1016/j.jviromet.2020.113925

3. Smithgall, M. C., Scherberkova, I., Whittier, S., & Green, D. A. (2020). Comparison of Cepheid Xpert Xpress and Abbott ID Now to Roche cobas for the Rapid Detection of SARS-CoV-2. Journal of Clinical Virology, 128, 104428. https://doi.org/10.1016/j.jcv.2020.104428

4. Lieberman, J. A., Pepper, G., Naccache, S. N., Huang, M.-L., Jerome, K. R., & Greninger, A. L. (2020). Comparison of Commercially Available and Laboratory-Developed Assays for In Vitro Detection of SARS-CoV-2 in Clinical Laboratories. Journal of Clinical Microbiology, 58(8), e00821–20. https://doi.org/10.1128/JCM.00821-20

5. Centers for Disease Control and Prevention (2022). COVID Data Tracker. US Department of Health and Human Services, CDC. https://covid.cdc.gov/covid-data-tracker

6. U.S. Food & Drug Administration. (2021). SARS-CoV-2 Viral Mutations: Impact on COVID-19 Tests. FDA. https://www.fda.gov/medical-devices/coronavirus-covid-19-and-medical-devices/sars-cov-2-viral-mutations-impact-covid-19-tests

7. Fox-Lewis, S., Fox-Lewis, A., Harrower, J., Chen, R., Wang, J., de Ligt, J., McAuliffe, G., Taylor, S., & Smit, E. (2021). Lack of N2-gene amplification on the Cepheid Xpert Xpress SARS-CoV-2 assay and potential novel causative mutations: A case series from Auckland, New Zealand. IDCases, 25, e01233. https://doi.org/10.1016/j.idcr.2021.e01233

8. Microbiologics. (2020). HE0065N Inactivated SARS-CoV-2 Whole Virus (Pellet) Instructions For Use. Microbiologics. https://www.microbiologics.com/core/media/media.nl?id=4532505&c=915960&h=cd311062442100c8242d&_xt=.pdf

9. Cepheid. (2021). Xpert® Xpress SARS-CoV-2 Instructions for Use. Cepheid. https://www.cepheid.com/Package%20Insert%20Files/Xpress-SARS-CoV-2/Xpert%20Xpress%20SARS-CoV-2%20Assay%20ENGLISH%20Package%20Insert%20302-3750%20Rev.%20F.pdf

10. Srivatsan, S., Heidl, S., Pfau, B., Martin, B. K., Han, P. D., Zhong, W., van Raay, K., McDermot, E., Opsahl, J., Gamboa, L., Smith, N., Truong, M., Cho, S., Barrow, K. A., Rich, L. M., Stone, J., Wolf, C. R., McCulloch, D. J., Kim, A. E., … Starita, L. M. (2022). SwabExpress: An End-to-End Protocol for Extraction-Free COVID-19 Testing. Clinical Chemistry, 68(1), 143–152. https://doi.org/10.1093/clinchem/hvab132

11. Bedford, T., Greninger, A. L., Roychoudhury, P., Starita, L. M., Famulare, M., Huang, M.-L., Nalla, A., Pepper, G., Reinhardt, A., Xie, H., Shrestha, L., Nguyen, T. N., Adler, A., Brandstetter, E., Cho, S., Giroux, D., Han, P. D., Fay, K., Frazar, C. D., … Jerome, K. R. (2020). Cryptic transmission of SARS-CoV-2 in Washington state. Science, 370(6516), 571–575. https://doi.org/10.1126/science.abc0523

12. Paredes, M. I., Lunn, S. M., Famulare, M., Frisbie, L. A., Painter, I., Burstein, R., Roychoudhury, P., Xie, H., Bakhash, S. A. M., Perez, R., Lukes, M., Ellis, S., Sathees, S., Mathias, P. C., Greninger, A., Starita, L. M., Frazar, C. D., Ryke, E., Zhong, W., … Oltean, H. N. (2022). Associations between SARS-CoV-2 variants and risk of COVID-19 hospitalization among confirmed cases in Washington State: A retrospective cohort study (p. 2021.09.29.21264272). medRxiv. https://doi.org/10.1101/2021.09.29.21264272

